# Onsite virtual classroom versus physical classroom: A comparative study of learning and student perception in orthopaedic training

**DOI:** 10.1101/2020.04.13.20063164

**Authors:** Kishore Puthezhath

## Abstract

**Introduction:** More students are posted in the orthopaedic out-patient departments than before. Lack of clinical space to accommodate students is a frequently cited problem. Virtual learning allows for an adjustable location and is scalable. We aimed to compare both the effectiveness and student satisfaction level between onsite virtual classroom and physical classroom in teaching orthopaedics to a group of undergraduate medical students.

**Methods:** A quasi-experimental non-equivalent group design study was conducted with 49 final-year medical students participating in orthopaedic training between November 2019 to January 2020. These students were randomly allocated into two groups, experimental (virtual classroom) and comparison (physical learning). The experimental group received an onsite virtual learning environment, whereas the control group received the same content in a physical classroom. Pre and post-tests that consisted of multiple choice questions were administered. At the end of the class, the students completed a 5-point Likert scale satisfaction level evaluation questionnaire.

**Results:** The post-test knowledge scores between virtual classroom (M=78.2,SD=12.74) and Physical classroom (M=77.92,SD=10.31) were not different (z= 0.00, p=1.00). Though the overall perceptions of learning were positive, some frustrations were apparent and the student satisfaction was significantly better (z=-4.60, p value=0.00) with the physical classroom (M=3.83,SD=0.16) compared to virtual classroom (M=3.5,SD=0.23).

**Conclusions:** Onsite virtual activities are not as satisfying as physical classroom in the opinion of the students, but they are successful strategies in learning that can be used in outpatient orthopaedic clinics to address the problem of lack of space. Students learn content focused on orthopaedic clinical learning objectives as well using onsite virtual classroom as they do in the traditional classroom setting.

## Introduction

In undergraduate medical education there is a trend away from ward-based teaching towards out-patient teaching. This may have developed as a necessary response to decreasing numbers of inpatients(1,2). With new demands on administrators in the delivery of medical school curriculum, more undergraduate students are posted in the department of Orthopaedics during a particular point of time. This has led to a frequently cited problem of lack of clinical space to accommodate students (1). Virtual learning allows for an adjustable location and is scalable.

Virtual classrooms (VCR) constitute a new promising structure in education of health care personnel. VCR uses the Internet to reach and deliver curricular content and allows learners to interact from any physical location(3). Virtual learning modalities include open source and for-profit software solutions. Technology type intended to be used for this study is an open source free software platform called Zoom(4,5).

Several studies have demonstrated that virtual classrooms are an efficient and effective way to enhance students’ learning. However, those studies tried to address the problem of how can the content-based curriculum be delivered to students at offsite locations, who are not able to attend regular lectures. A literature review on 26.9.19 using Papers 3 for Macintosh, using the keywords “Virtual Classroom” and “interactive video conferencing” retrieved no study comparing virtual classroom with physical classroom for onsite undergraduate medical education.

We aimed to explore the knowledge and the satisfaction levels between virtual classroom training and physical classroom after the completion of synchronous orthopaedic classes. We hypothesized that the setting of the class (Virtual or Physical) does not affect the knowledge and the satisfaction levels.

## Methods

### Study Design and setting

This was a quasi-experimental study with a non-equivalent control group design conducted at a Medical college at Kerala University of Health Sciences, India. The Virtual classroom system used in the study was the freeware version of the video communication software, Zoom. Physical classes taken from the Orthopaedic out-patient clinic were synchronously streamed to the group of students attending the virtual class using an iPad Pro. Students in the virtual group were seated in a classroom near the out-patient clinic and received the class using the Zoom application for mobile devices.

### Study Participants

Participants in two groups comprised final-year medical students participating undergraduate training between November 2019 to January 2020 during their rotation through orthopaedics. There were 3 rotations with approximately 17 students per rotation. 54 ninth semester MBBS students were enrolled; 49 students were available for the study. They were randomly grouped into experimental and control by taking a lot written A(Virtual) or B(Physical) respectively. None of the students in either group had been exposed to virtual classroom previously. The analyses were done with 25 students in the virtual classroom group and 24 participants in the physical group.

The Institutional Ethics Committee of the Medical College Hospital approved the study. Written informed consent was obtained from the participating students because the study involved modification of existing curriculum delivery method in an educational setting. The study was conducted according to the Belmont report ethical considerations: all participants’ data were confidential, no harm would be afflicted upon participants during the study, and their refusal in doing tests or questionnaires in the study would have no impact on their course assessment or grades.

### Assessment and survey

Group A (the experimental,VCR group) was given three onsite clinical tutorials virtually using Zoom. For the second control group B, in-person tutorials were conducted during the same period. Total number of interventions were 6 (Six). For the VCR group, there were 25 students assigned to 3 VCR groups; each VCR group consisted of 7-8 students. Each VCR session was 1 h per clinical session and was facilitated by a single faculty member. The 24 students assigned to the 3 PCR groups received the same class from the out-patient clinic synchronously for 1 h per session as that of the VCR group.

The faculty member then wrapped up the class and answered any final questions the students may have had. Immediately before and after the intervention, students were assessed for the recall in the topics covered using MCQs with thatquiz online software(6). Qualitative evaluation of students’ experience was done by online Likert questionnaires (Using Poll everywhere software)(7). Crossover was done after the post test to address possible ethical issues.

## Data Analysis

Descriptive statistics were generated from the test scores and attitude scales. Data were presented as mean and standard deviation (SD). The Wilcoxon test was used to compare pretest and post test learning scores. Mann Whitney U test was used to compare differences between the VCR and PCR groups, without controlling for baseline differences in the abilities, because the baseline differences (Pretest score) in the abilities between groups were statistically insignificant. PSPP 1.2.0-2 for MacOS was used to analyze the data(8) .

## Results

There were more female participants (63%) than males (37%) (Figure 1). To compare the baseline differences in the abilities between the two groups, Mann Whitney U test, using pretest scores as the variable was used. Though there was no statistically significant (p=0.6) between the pretest scores of VCR and PCR groups, the box plot showed the scores to be spread over a larger area (20-100) in PCR group when compared to VCR group (0-90) (Figure 2). This might have been balanced by an outlier with a score of zero in the VCR group. The pretest scores of both males and females showed no statistically significant difference, with a median score of 40% (Figure 3). The post test knowledge scores of both VCR and PCR group were significantly improved to 80 when compared to pretest scores. However, at the end of the class, the Post test knowledge scores between virtual classroom (M=78.2,SD=12.74) and Physical classroom (M=77.92,SD=10.31) were not different (z= 0.00, p=1.00). (Figure 4). Interestingly, the spread of the box plot for the post test of VCR group (55-100) was more when compared to PCR group (60-90), the significance of which is less apparent to me. There was one outlier with a 50% mark in PCR group. Post test scores showed no statistically significant difference between males and females, though females had a lower score (Figure 5). Though the overall perceptions of learning were positive the student satisfaction was significantly better (z=-4.60, p value=0.00) with the physical classroom (M=3.83,SD=0.16) compared to virtual classroom (M=3.5,SD=0.23) (Figure 6).

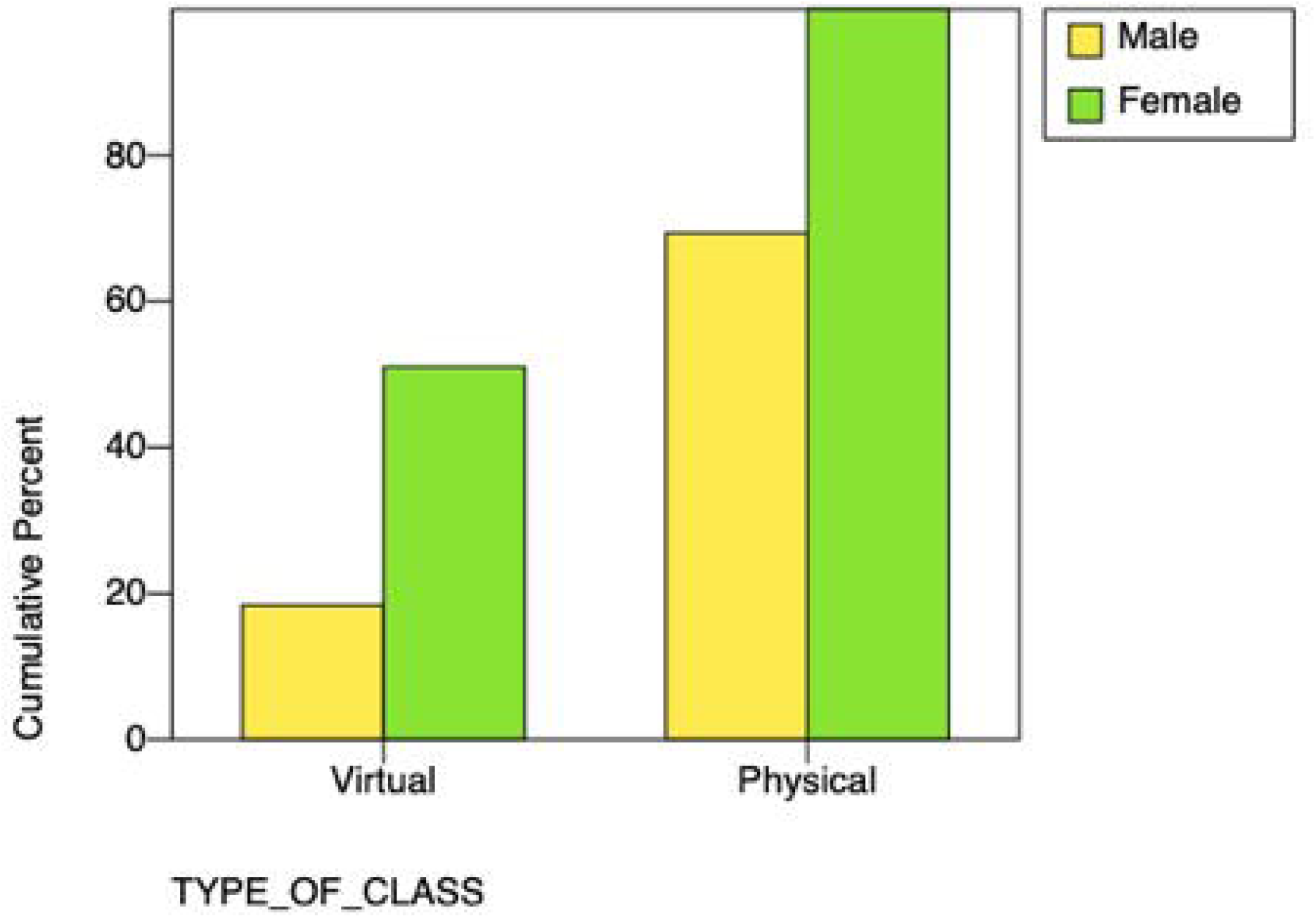

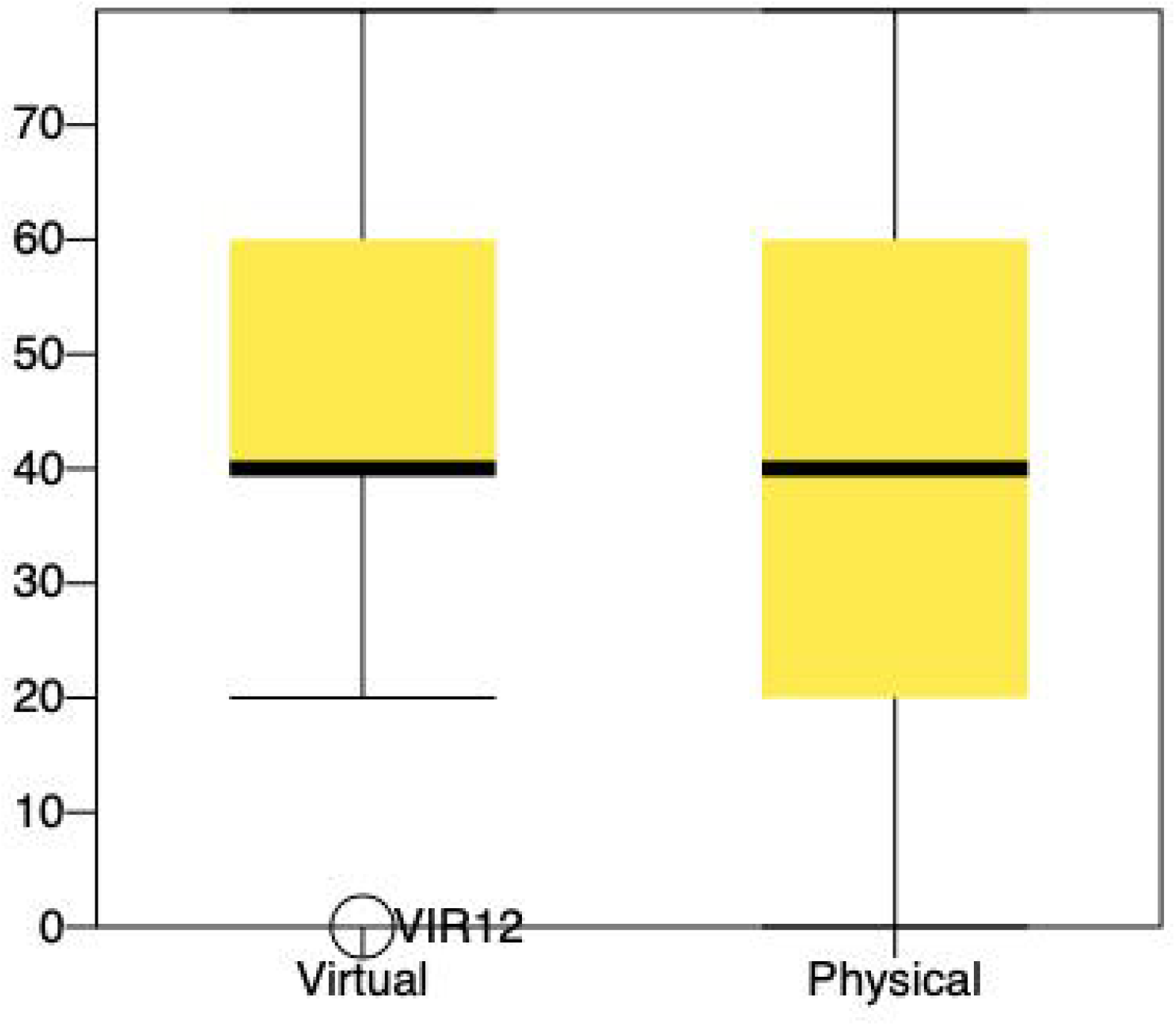

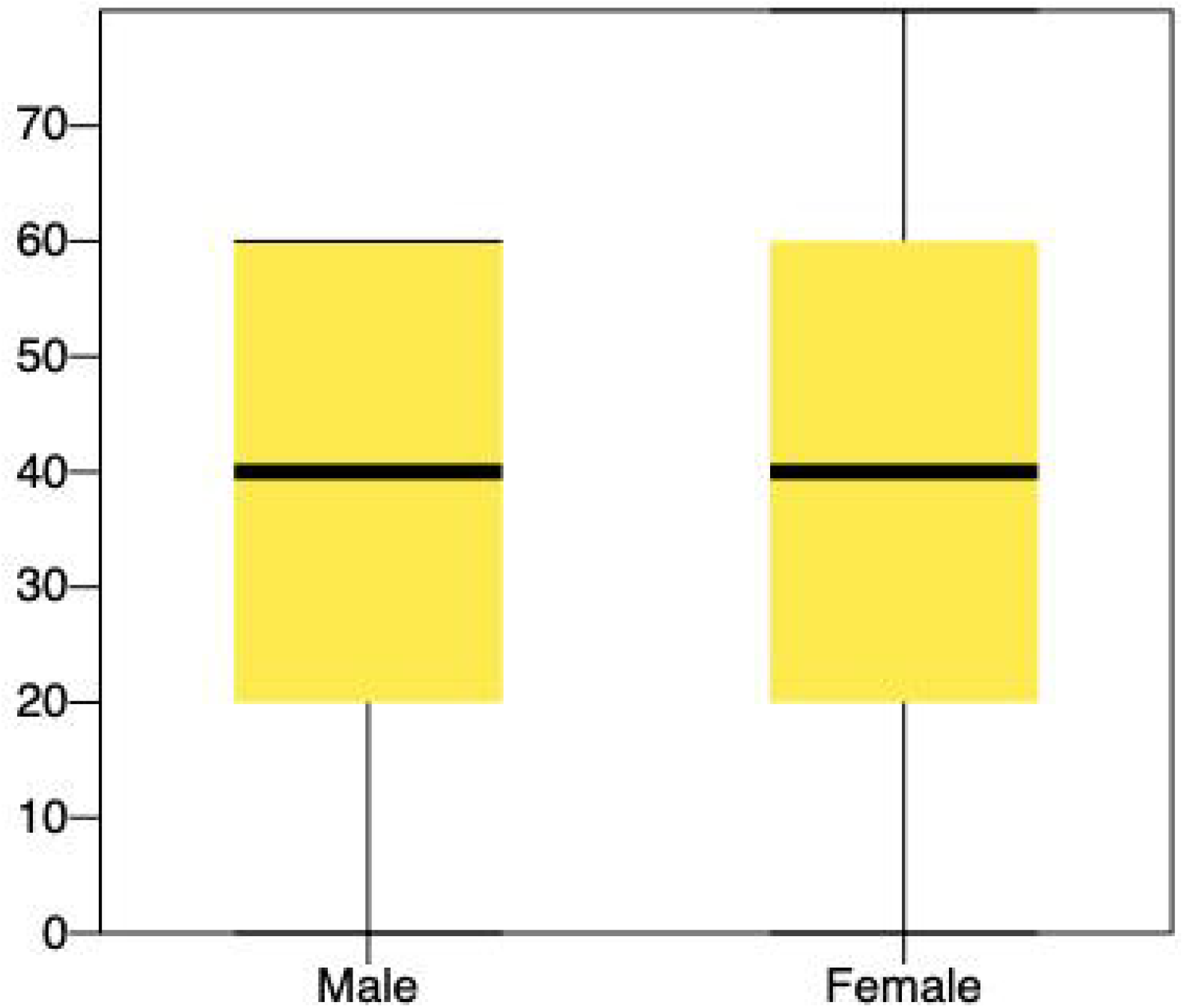

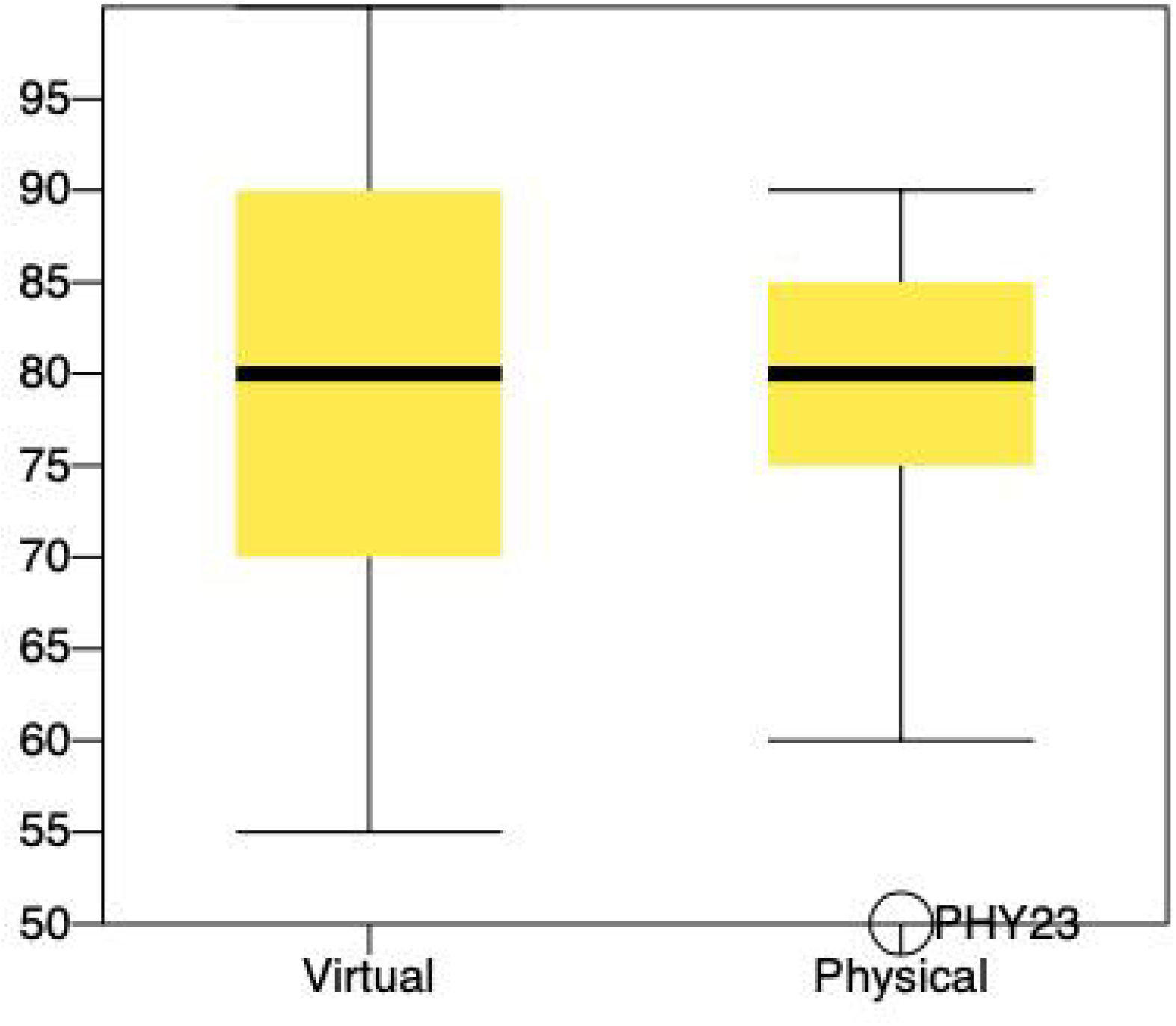

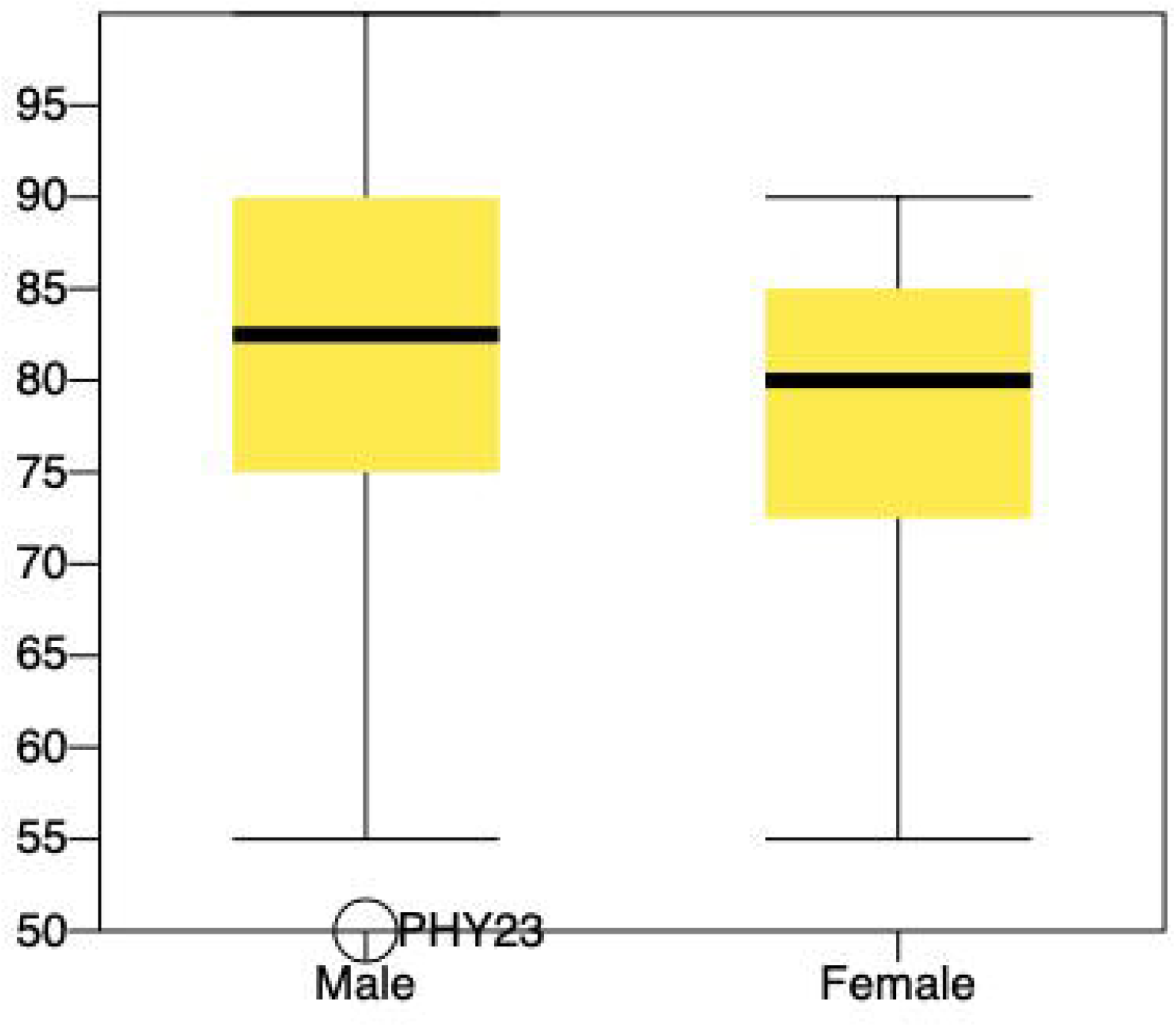

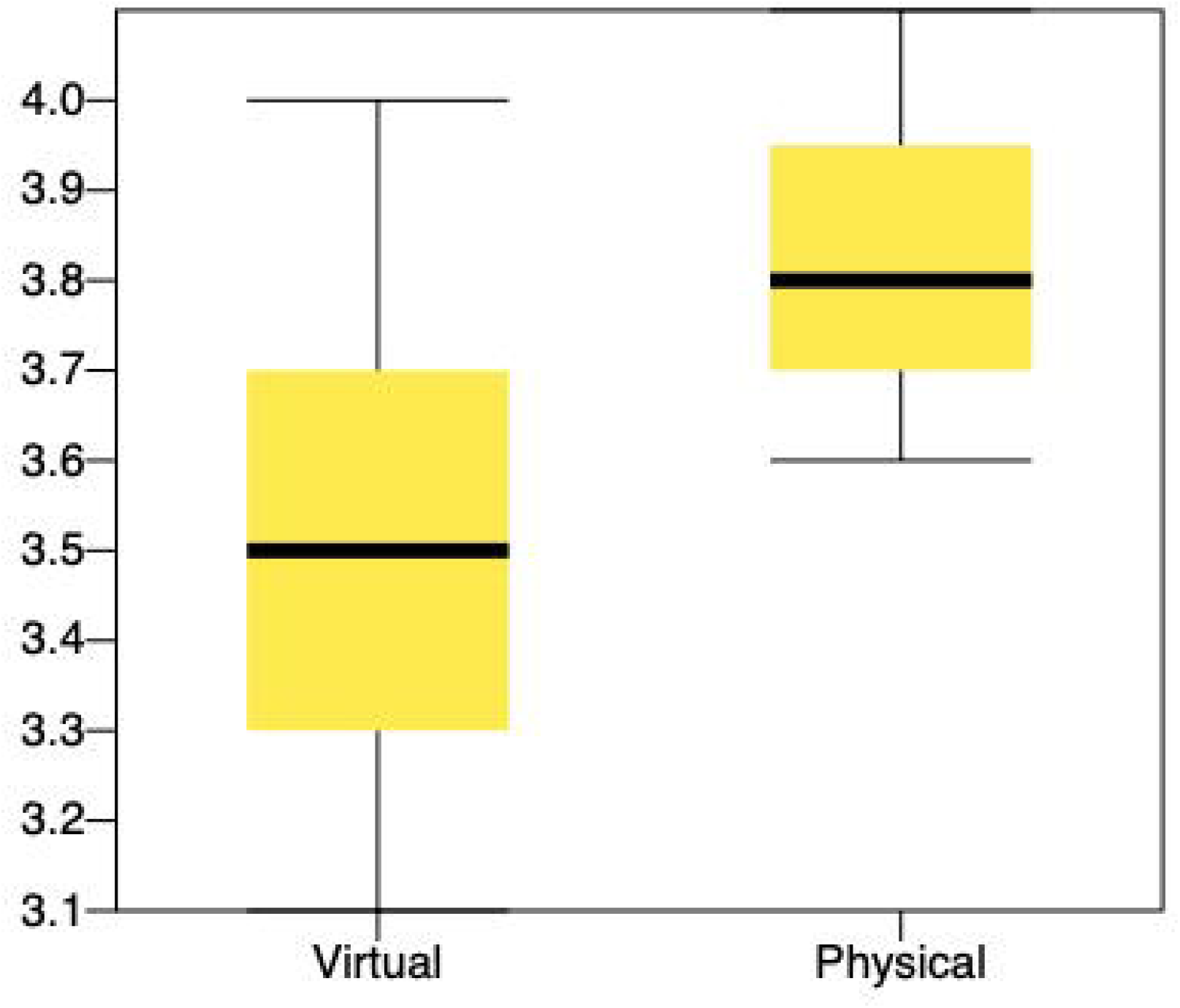

## Discussion

The development of web-based virtual classroom historically rises from organizational constraints such as repetitive student’s clinical rotations in hospitals that are geographically disparate from the main educational campus or to address the high rate of absenteeism (9)(10). Hence, although multiple studies suggest that technology-mediated learning can be used for effective clinical learning, those studies were conducted at off-site locations(11). One such offsite study evaluating the effectiveness of live lectures delivered via video-conferencing with that of in-person lectures reported no statistically significant difference in the performance of medical students who attended the national licensing examination(12).

In the study that I describe in this report, the students had also received lectures via virtual classroom, but at on-site and this was to address the well known problem of lack of clinic space to accommodate more students. The solution proposed by this study is cost effective and scalable, unlike the proposal by Feltovich et al. to develop ambulatory sites for teaching other than hospital out-patient clinics, for example, private offices, community clinics, and health maintenance organisations to resolve the limitations(1). My study supports the concept that interactive virtual classrooms can be an effective tool to teach orthopaedic objectives to undergraduate medical students. This method of instruction was as successful as traditional classroom instruction in achieving learning objectives as measured by the Post test.

When teaching students new information, it is important to consider how this information is delivered as it is necessary to encourage student engagement and satisfaction. A decrease in satisfaction may impact on their engagement and acquisition of knowledge, including their ability to recall and retain information(10).

Callas and Bertsch had shown that the level of satisfaction among the Medical students of lectures attended from rural sites via interactive videoconferencing was high for most aspects of remote lecture attendance, although not quite as high as for in-person attendance. In the present study, the student satisfaction was significantly better with physical classroom compared to virtual classroom(12,13).

This could be due to several factors. Despite its advantages, learning in a virtual environment sometimes creates technical issues, and leads to student’s isolation and decreased Interaction is often decreased in virtual learning methods(9). In addition, because technical problems such as dropped calls or inadequate audio or visual clarity can have a negative impact on remote viewers. Increasing system reliability and quality is a necessity for implementing a satisfactory virtual classroom. However, in my study, the engagement and the ability to recall information were similar in both groups.

## Conclusions

Onsite virtual activities are not as satisfying as physical classroom in the opinion of the students, but they are successful strategies in learning that can be used in outpatient orthopaedic clinics to address the problem of lack of space. Students learn content focused on orthopaedic clinical learning objectives as well using onsite virtual classroom as they do in the traditional classroom setting.

## Data Availability

Master chart and data analysis output is available with the author who can share it at any time.

## Notes

### Competing Interest Statement

The authors have declared no competing interest.

### Summary of Updates

A typing error which lead to wrong marking of the groups

